# Behavioural outcomes following in utero exposure to anti-seizure medication: protocol for a systematic review

**DOI:** 10.1101/2022.08.22.22278046

**Authors:** Eliza Honybun, Emily Cockle, Charles B Malpas, Terence J. O’Brien, Frank J.E. Vajda, Piero Perucca, Genevieve Rayner

## Abstract

**Introduction:** Prenatal exposure to certain antiseizure medications (ASMs) has been associated with increased risk of adverse neurodevelopmental outcomes in offspring. While the cognitive and intellectual outcomes of ASM-exposed offspring have been well-described, the long-term behavioural and functional sequalae in these children have received less attention. This systematic review aims to synthesise evidence on the relationship between prenatal ASM exposure and postnatal adverse neurodevelopmental outcomes, focusing on non-cognitive and intellectual domains of neurodevelopment including reduced social, emotional, behavioural, and adaptive functioning, as well as the frequency of neurodevelopmental and psychiatric disorders. This will have meaningful clinical implications for how we counsel women taking ASMs in pregnancy.

**Methods and analysis:** Studies reporting predefined neurodevelopmental outcomes will be identified by electronic searches of MEDLINE, PsychINFO, EMBASE, as well as additional manual and grey literature searches. Eligible studies will report outcomes of offspring exposed to ASMs in utero either prospectively or retrospectively from 1990 to present, with screening performed in duplicate. We will use the Newcastle-Ottawa Scale to conduct methodological quality assessments of included observational studies. A narrative synthesis will be used to report on the review findings. Meta-analysis is not anticipated.

**Ethics and dissemination:** Ethics clearance is not required for the current study. The systematic review will be prepared as a journal article and published in a peer-reviewed journal upon completion.

**PROSPERO registration number:** PROSPERO CRD42021281919

**Article Summary:** Strengths and limitations of this study

- This protocol was developed and written according to the PRISMA-P guidelines
- Publication of this protocol ensures transparency and reproducibility of the methods of the systematic review, as well as reduces the likelihood of review duplication
- Restricting publications to English only may introduce bias whereby some relevant data is not included
- Meta-analysis is not likely to be possible due to heterogeneity in study methodology, reducing the strength of the conclusions that can be drawn
- Targeting psychosocial and behavioural outcomes allows for a more nuanced understanding of the long-term clinical consequences of prenatal ASM-exposure

## Introduction

The use of certain antiseizure medications (ASMs) during pregnancy is associated with an increased risk of various adverse neurodevelopmental outcomes in exposed offspring [1, 2].Neurodevelopment is an extensive process that underpins the growth and maturation of the central nervous system, including brain pathways that support biological, cognitive, behavioural, psychological, and adaptive aspects of human functioning[3, 4]. Many studies investigating the neurodevelopment sequelae of in utero exposure to ASMs have focused heavily on documenting global intellectual ability, such as IQ, with more nuanced clinical outcomes such as adaptive, behavioural, and psychological dysfunction often overlooked.

While several studies have aimed specifically to characterise the clinical and behavioural outcomes following prenatal ASM exposure[5-8], a wealth of clinical and psychometric data also exist within larger neurodevelopmental studies, that are seldom scrutinised. This study aims to systematically review literature describing the neurodevelopmental outcomes of ASM-exposed offspring, focusing exclusively on non-cognitive outcomes such as adaptive, behavioural, social, and psychological outcomes for the purposes of outlining a ‘clinical phenotype’ of prenatal ASM exposure.

## Methods and analysis

### Protocol and registration

This systematic review protocol has been developed according to the Preferred Reporting Items for Systematic review and Meta-Analysis Protocols (PRISMA-P) guidelines[9]. The review protocol was registered on PROSPERO on 29 October 2021 by E.H. (ID CRD42021281919).

### Intervention assessed

The intervention assessed will be in-utero exposure to ASMs.

### Control populations

A control or comparator group is not required.

### Outcome measures

The review will target the following behavioural outcomes:

1. Social development: the ability of a child to communicate and interact with those around them, which can be psychometrically measured by instruments such as the Social Skills Rating Scale (SSRS)[10] and Child & Adolescent Social Cognitions Questionnaire (CASCQ)[11],
2. Adaptive functioning: practical skills and behaviours that are learned in order to function within one’s environment (e.g., personal grooming, helping with chores, engaging in leisure activities), which can be measured by parental rating of daily living skills using tools like the Vineland[12] and Adaptive Behaviour Assessment System (ABAS)[13],
3. Emotional functioning: the awareness, expression, and regulation of emotions, which can be measured by the Behaviour Assessment System for Children (BASC)[14] and Child Behaviour Checklist (CBCL)[15],
4. Formal neurodevelopmental/neuropsychiatric disorder or syndrome (e.g., ASD, ADHD, conduct disorder, anxiety, depression)

### Literature search

Electronic searches of MEDLINE, PsychINFO, and EMBASE will be conducted using a set of pre-defined search criteria. Key search terms will include variations or synonyms of the following: pregnancy (“pregnancy” or “prenatal” or “in-utero”), anti-seizure medication (“antiepileptic mediation”, “AED”, “anticonvulsant”), neurodevelopmental outcomes (“neurodevelopment”, “autism”, “attention deficit hyperactivity disorder”). The search strategy will apply filters for human studies, English language, and studies published after 1990. Grey literature searches will be conducted, as well as additional searches of Google Scholar (related articles) and article reference lists to identify additional articles that were not retrieved from the primary electronic searches.

### Selection criteria

#### Inclusion criteria

This systematic review will consist of articles that study offspring born to mothers who took ASMs at any time during pregnancy. We will include ASM monotherapy and polytherapy exposures, and there will be no restrictions on the indication for ASM usage. The mothers of these offspring will be included in the review to examine the contribution of relevant demographic factors such as maternal age and IQ to offspring neurodevelopmental outcomes. The following study designs will be included in the review: 1) observational cohort studies, 2) population-based datasets, 3) register-based studies, 4) case-control studies. Eligible studies will report original data pertaining to behavioural outcomes following prenatal ASM exposure, be published after the year 1990 and be available in English.

#### Exclusion criteria

Animal studies and studies that do not report original data (i.e., review studies, letters, editorials, and commentaries) will be excluded. Case reports or studies with fewer than 10 participants will be excluded. Studies that only report on anatomical or pregnancy outcomes (e.g., major congenital malformations, neonatal birth weight, stillbirth) will be excluded, as will studies that only report superordinate cognitive outcomes (i.e., intelligence quotient) in the absence of other clinical or psychometric data relevant to neurodevelopmental outcomes. We will exclude studies where the full text is not available after requests to the authors.

### Selection of studies for inclusion in the review

Two reviewers will independently undertake title and abstract screening for studies that meet the including criteria using Covidence, an online software program for conducting systematic reviews[16]. Full texts of relevant studies will be obtained and will undergo a full-text review to confirm eligibility for inclusion. Any conflicts will be resolved by a third reviewer.

### Data collection

Two reviewers will independently extract information from the included studies using a pre-defined extraction form within the Covidence software. Extracted data pertaining to study and population characteristics will include study design, country, recruitment method, ASM exposure (including ASM type, daily dosage, use as monotherapy or in combination therapy), sample sizes of exposed offspring, offspring demographic information (mean age at follow-up, sex), psychometric instruments, duration of follow up (if applicable). Extracted outcome data will pertain to neurodevelopmental outcomes; specifically, social, emotional, adaptive functioning, and diagnosis of neurodevelopmental and/or psychiatric disorders. Where applicable, prevalence ratios, mean differences, and relative risks will be reported with accompanying p values and confidence intervals.

### Risk of bias assessment

Two authors will independently conduct quality assessments of all included studies in the review using the Newcastle Ottawa Scale (NOS)[17]. The NOS is a risk-of-bias tool recommended for evaluation of study quality for non-randomised studies. Studies are judged against the categories of A) participant selection; B) comparability of study groups; C) exposure; and D) outcome. The risk of bias score will be reported for each separate domain, and any disagreements will be resolved through consensus.

### Data synthesis

This study will utilise a narrative review to synthesise the social, emotional, behavioural, adaptive, and neurodevelopmental outcomes to describe a clinical phenotype of offspring exposed to ASMs in utero. Findings will be separated by ASM type to create a ‘profile’ for each ASM, with monotherapy and polytherapy findings reported separately where appropriate. Where there are insufficient data for specific ASMs, the review will highlight the need for further research. Within the section for each ASM, the quality of studies will also be commented on to evaluate the validity of the findings.

Due to the clinical nature of the study sample, it is anticipated that there will be a small number of eligible studies, with differences in methodology and outcome measures further limiting the scope for meta-analysis[18].

### Patient and public involvement

Patients or the public were not involved in the design, or conduct, or reporting, or dissemination plans of this research.

### Ethics and dissemination

As the current study is based on already published data, ethics clearance is not required. The systematic review will be written into a journal article and published in a peer-reviewed journal, with raw data available upon request. The systematic review will be reported in accordance with PRISMA guidelines.

## Discussion

A comprehensive understanding of the neurodevelopmental sequelae of prenatal ASM exposure is crucial to advance knowledge of the teratogenic effects of ASMs. Research to date in this area has focused predominantly on broad cognitive outcomes like IQ score[19-21], rather than more nuanced behavioural and adaptive outcomes. To this end, there is a need for a systematic review to expand this framework to include other domains of neurodevelopment, such as social, emotional, psychological, behavioural, and adaptive outcomes that are known to have major impacts of the child’s attainment of age-appropriate educational, vocational, and personal milestones. Understanding the clinical phenotype of ASM-exposed offspring will provide greater context and refinement to pre-existing cognitive research and provide clear delineation of the functional consequences of ASM usage on offspring during pregnancy.

We anticipate that there will be few studies focused specifically on documenting behavioural, rather than cognitive outcomes following prenatal ASM exposure, but expect to find many clinical instruments within studies investigating cognitive and intellectual outcomes. To the best of our knowledge, there are currently no other systematic reviews that solely consider behavioural and psychosocial outcomes of prenatal ASM exposure, however, this review will complement pre-existing systematic reviews and meta-analyses on other neurodevelopmental outcomes[1, 22].This review will report and synthesise findings across a range of sources, including published reports and grey literature, to pool together behavioural data that can be under-acknowledged in the context of significant cognitive data with large effect sizes.

This systematic review will help to further inform pre-conception counselling of women taking ASMs, but also enhance the paediatric monitoring of ASM-exposed offspring. Understanding the types of problems ASM-exposed offspring are likely to present with will identify areas for intervention and allow for any deficits to be remediated early, which can lead to improvements in functional and academic outcomes.

## Data Availability

The systematic review will be written into a journal article and published in a peer-reviewed journal, with raw data available upon request.

## Author contributions

EH conceived and designed the systematic review. EH, GR, CM, PP, FV, and TOB contributed to the conception and development of the study protocol. EH drafted the protocol and registered it on PROSPERO. EH is the first author and corresponding author of the review.

## Acknowledgements

The authors are grateful for the support from Dr. Ana Antonic-Baker in the conceptualisation and development of the review protocol.

## References

1. Bromley, R., et al., Treatment for epilepsy in pregnancy: neurodevelopmental outcomes in the child. Cochrane Database of Systematic Reviews, 2014(10).

2. Meador, K.J., et al., Two-Year-Old Cognitive Outcomes in Children of Pregnant Women with Epilepsy in the Maternal Outcomes and Neurodevelopmental Effects of Antiepileptic Drugs Study. JAMA Neurology, 2021: p. 158.

3. Siegler, R.S., et al., How children develop. Fifth edition. ed. 2017: Worth Publishers, Macmillan Learning.

4. Anderson, V., E. Northam, and J. Wrennall, Developmental neuropsychology: A clinical approach. 2018: Routledge.

5. Huber-Mollema, Y., et al., Behavioral problems in children of mothers with epilepsy prenatally exposed to valproate, carbamazepine, lamotrigine, or levetiracetam monotherapy. Epilepsia, 2019. 60(6): p. 1069–1082.

6. Deshmukh, U., et al., Behavioral outcomes in children exposed prenatally to lamotrigine, valproate, or carbamazepine. Neurotoxicol Teratol, 2016. 54: p. 5–14.

7. Richards, N., et al., Developmental outcomes at age four following maternal antiepileptic drug use. Epilepsy Behav, 2019. 9: p. 7–79.

8. Vinten, J., et al., The behavioral consequences of exposure to antiepileptic drugs in utero. Epilepsy and Behavior, 2009. 14(1): p. 197–201.

9. Moher, D., et al., Preferred reporting items for systematic review and meta-analysis protocols (PRISMA-P) 2015 statement. Systematic reviews, 2015. 4(1): p. 1–9.

10. Elliott, S.N. and F.M. Gresham, Social skills rating system. American Guidance Service: Circle pines, MN, USA, 1990.

11. Leigh, E. and D.M. Clark, Development and Preliminary Validation of the Child & Adolescent Social Cognitions Questionnaire. Child Psychiatry & Human Development, 2021.

12. Sparrow, S.S., D.V. Cicchetti, and C.A. Saulnier, Vineland adaptive behavior scales. Bloomington, MN: NCS Pearson. 2016, Inc.

13. Harrison, P.L. and T. Oakland, Adaptive behavior assessment system. 2000: Psychological Corporation San Antonio, TX.

14. Reynolds, C.R., Behavior assessment system for children. The Corsini encyclopedia of psychology, 2010: p. 1–2.

15. Achenbach, T.M., The Child Behavior Checklist and related instruments. 1999.

16. Covidence, Covidence systematic review software, Veritas Health Innovation, Melbourne, Australia.

17. Wells GA S.B., O’Connell D, Peterson J, Welch V, Losos M, et al., The Newcastle-Ottawa Scale (NOS) for assessing the quality of nonrandomised studies in meta-analyses. 2014.

18. Deeks, J.J., et al., Analysing data and undertaking meta-analyses. Cochrane handbook for systematic reviews of interventions, 2019: p. 241–284.

19. Baker, G., et al., School aged IQ in children prenatally exposed to antiepileptic drugs. Epilepsia, 201. 54(SUPPL.): p. 7–8.

20. Nadebaum, C., et al., The australian brain and cognition and antiepileptic drugs study: Iq in school-aged children exposed to sodium valproate and polytherapy. Journal of the International Neuropsychological Society, 2011. 17(1): p. 1–142.

21. Meador, K.J., et al., Relationship of child IQ to parental IQ and education in children with fetal antiepileptic drug exposure. Epilepsy & behavior : E&B, 2011. 21(2): p. 147–52.

22. Knight, R., A. Wittkowski, and R.L. Bromley, Neurodevelopmental outcomes in children exposed to newer antiseizure medications: A systematic review. Epilepsia, 2021.

